# Development of a deep learning classifier to accurately distinguish COVID-19 from look-a-like pathology on lung ultrasound

**DOI:** 10.1101/2020.10.13.20212258

**Authors:** Robert Arntfield, Blake VanBerlo, Thamer Alaifan, Nathan Phelps, Matt White, Rushil Chaudhary, Jordan Ho, Derek Wu

## Abstract

**Objectives:** Lung ultrasound (LUS) is a portable, low cost respiratory imaging tool but is challenged by user dependence and lack of diagnostic specificity. It is unknown whether the advantages of LUS implementation could be paired with deep learning techniques to match or exceed human-level, diagnostic specificity among similar appearing, pathological LUS images.

**Design:** A convolutional neural network was trained on LUS images with B lines of different etiologies. CNN diagnostic performance, as validated using a 10% data holdback set was compared to surveyed LUS-competent physicians.

**Setting:** Two tertiary Canadian hospitals.

**Participants:** 600 LUS videos (121,381 frames) of B lines from 243 distinct patients with either 1) COVID-19, Non-COVID acute respiratory distress syndrome (NCOVID) and 3) Hydrostatic pulmonary edema (HPE).

**Results:** The trained CNN performance on the independent dataset showed an ability to discriminate between COVID (AUC 1.0), NCOVID (AUC 0.934) and HPE (AUC 1.0) pathologies. This was significantly better than physician ability (AUCs of 0.697, 0.704, 0.967 for the COVID, NCOVID and HPE classes, respectively), p < 0.01.

**Conclusions:** A deep learning model can distinguish similar appearing LUS pathology, including COVID-19, that cannot be distinguished by humans. The performance gap between humans and the model suggests that subvisible biomarkers within ultrasound images could exist and multi-center research is merited.

## Introduction

Lung ultrasound (LUS) is an imaging technique deployed by clinicians at the point-of-care to aid in the diagnosis and management of acute respiratory failure. With accuracy matching or exceeding chest X-ray (CXR) for most acute respiratory illnesses,^1–3^ LUS additionally lacks the radiation and laborious workflow of computed tomography (CT). Further, as a low cost, battery operated modality, LUS can be delivered at large scale in any environment and is ideally suited for pandemic conditions.^4^

B lines are the characteristic pathological feature on LUS, created by either pulmonary edema or non-cardiac causes of interstitial syndromes. The latter includes a broad list of conditions ranging from pneumonia, pneumonitis, acute respiratory distress syndrome (ARDS) or fibrosis.^5^ While an accompanying thick pleural line is helpful in differentiating cardiogenic from non-cardiogenic causes of B lines,^6^ reliable methods to differentiate non-cardiogenic causes from one another on LUS have not been established. Additionally, user dependent interpretation of LUS contributes to wide variation in disease classification,^7,8^ creating urgency for techniques that improve diagnostic precision and reducing user-dependence.

Deep learning (DL), a foundational strategy within present-day artificial intelligence (AI) techniques, has been shown to meet or exceed clinician performance across most visual fields of medicine.^9–11^ Without cognitive bias or reliance on spatial relationships between pixels, DL ingests images as numeric sequences and evaluates for quantitative patterns that may reveal information that is unavailable to human analysis.^12^ With CT and CXR research maturing, ^13–15^ LUS remains comparably understudied with DL due to a paucity of organized, well labelled LUS data sets and the seeming lack of rich information in its minimalistic, artifact-based images.

In this study, we trained a neural network using LUS images of B lines from 3 different etiologies (hydrostatic pulmonary edema (HPE), ARDS and COVID-19). Using LUS-fluent physicians as comparison, we sought to determine if subvisible features in LUS images are available to a DL model that would allow it to exceed human limits of interpretation.

## Methods

### Data identification, extraction and labelling

After University of Western Ontario Research Ethics Board (REB 115723) approval, LUS exams performed at London Health Sciences Centre’s 2 tertiary hospitals were identified within our database of over 100,000 point-of-care ultrasound exams. The curation and oversight of this archive have previously been described.^16^ The goal of this study was to determine if a deep neural network could distinguish between the B line profiles of 3 different disease profiles, namely 1) hydrostatic pulmonary edema (HPE); 2) non-COVID ARDS (NCOVID) causes; and 3) COVID-19 ARDS (COVID). These profiles were chosen deliberately to challenge the neural network to classify images with obvious qualitative differences (HPE vs ARDS) and with no obvious differences (NCOVID vs COVID) between their B lines patterns (Figure 1, Videos 1, 2, 3). The COVID class consisted of confirmed cases of COVID-19 via reverse-transcriptase polymerase chain reaction test. The NCOVID class consisted of an assortment of causes: aspiration, community acquired pneumonia, hospital acquired pneumonia and viral pneumonias. Exams were conducted as part of patient encounters in the emergency department, intensive care unit and medical wards across the 2 hospitals.

**Figure 1:**
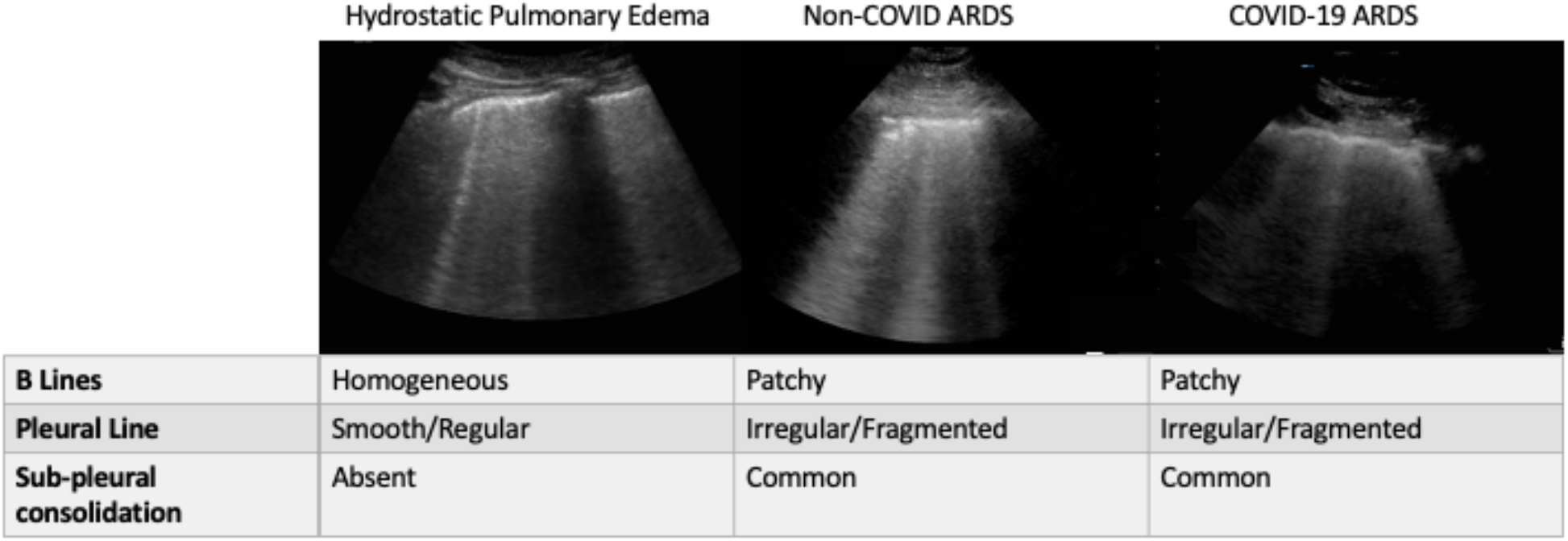
Sample images and lung ultrasound characteristics typical of the 3 lung pathologies that are the subject of our deep learning classifier (videos available in e-supplement as Videos 1, 2, 3).

Candidate exams for inclusion were identified using a sequential search by 2 critical care physicians, ultrasound experts (RA, TA) from within the finalized clinical reports of our database of LUS cases (Figure 2).

**Figure 2:**
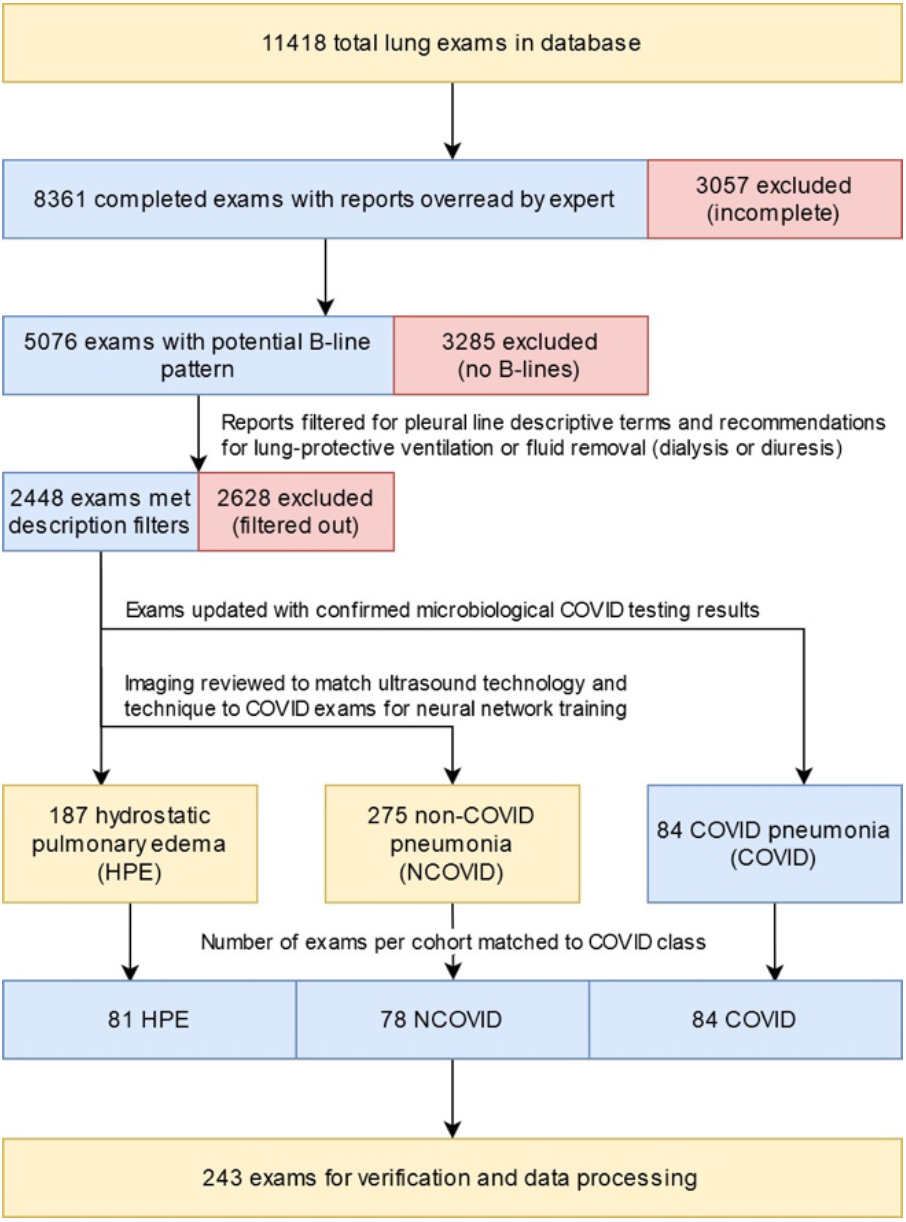
Data acquisition, selection and verification workflow.

Videos from our dataset represented a variety of ultrasound systems with phased array probe predominantly used for acquisition. Videos of the costophrenic region (which included solid abdominal organs, diaphragm, or other pleural pathologies such as effusions or trans-lobar consolidations) were excluded as 1) these regions did not contribute greatly to alveolar diagnoses, 2) this would introduce heterogeneity into the still image data, and 3) a trained clinician can easily distinguish between these pathologies and B lines. Duplicate studies were discarded to avoid overfitting. From each encounter, de-identified mp4 loops of B lines, ranging from 3-6 seconds in length with a frame rate ranging from 30-60/second (depending on the ultrasound system), were extracted. As COVID was the newest class available to our database, its comparably smaller number of encounters governed the number of encounters we extracted from HPE and NCOVID. A balanced volume of data for each class of image is important to avoid model over training on a single image class and/or overfitting.

### Data Preprocessing

The images used to train the model were all frames from the extracted LUS clips. Hereafter, a *clip* refers to a LUS video that consists of several *frames*. An *encounter* is considered to be a set of one or more clips that were acquired during the same LUS examination.

Preprocessing of each frame consisted of a conversion to grayscale followed by a script written by one of our team (JH) to scrub the image of extraneous information (index marks, logos, and manufacturer-specific user interface). See Supplementary Appendix for full details.

Data augmentation techniques were applied to images to each batch of training data during training experiments to combat overfitting. Augmentation transformations included random zooming in/out by ≤10%, horizontal flipping, horizontal stretching/contracting by ≤20%, vertical stretching/contracting (≤5%), and bi-directional rotation by ≤10°.

### Model Architecture and Training

In choosing an optimal architecture for our model, we investigated training from scratch on custom implementation of feedforward CNNs, residual CNNs as well as transfer learning methods.^17^ Ultimately, Xception architecture^18^ achieved the highest performance among the custom and 7 common architectures evaluated.

Individual preprocessed images were fed into the network as a tensor with dimensions 600×600×3. Although the images were originally greyscale, they were converted to RGB representation to ensure that the model input shape was compatible with the pre-trained weights. The output tensor of the final convolutional layer of the Xception model was subject to 2D global average pooling, resulting in a 1-dimensional tensor. Dropout at a rate of 0.6 was applied to introduce heavy regularization to the model and provided a noticeable reduction in overfitting. The final layer was a 3-node fully connected layer with softmax activation. The output of the model represents the probabilities that the model assigned to each of the 3 classes, all summing to 1.0. The argmax of this probability distribution was considered to be the model’s decision. To further combat overfitting, early stopping was applied by halting training if the loss on the validation set did not decrease over the most recent 15 epochs.^20^

For additional details on model selection, training, coding practice, our GitHub repository and hardware used in this project, please see Supplementary Appendix.

### Validation Strategy

A modification of the holdout validation method was used to ensure that the model selection process was independent of the model validation. Our holdout approach began with an initial split that randomly partitioned all encounters into a training set and 2 test sets (henceforth referred to as test-1 set and test-2 set). The distribution of encounters and frames after this split are shown in Table 1. It must be emphasized that all splits were conducted at the encounter level, not across all frames, thus ensuring that frames from the same clip did not appear in more than one partition. Test-1 was used to evaluate all of the candidate models so that a final model architecture and set of hyperparameters could be chosen. Test-2 was considered to be the holdout set, since it remained untouched throughout the model selection process and was only used during the final validation phase. A full account of validation methods can be found in the Supplementary Appendix.

**Table 1:** Distribution of clips and images assigned to each dataset

| Data Split | Encounters [% of total] | Frames [% of total] | Clips [% of total] |
| --- | --- | --- | --- |
| Training set | 204 | 99471 | 500 |
| Test-1 set | 19 (7.82%) | 9540 (7.86%) | 49 [8.00%] |
| Test-2 set | 20 (8.23%) | 12370 (10.19%) | 63 [10.29%] |

### Measuring Model Performance

The final model performance was determined by its results on our hold-back, independent dataset (test-2). The results were analyzed both at the individual frame level and at the encounter level. The latter was achieved through averaging the classifier’s predicted probabilities across all images from within that encounter. We assessed the model’s performance by calculating the area under the receiver operating characteristic curve (AUC), analyzing a confusion matrix, and calculating metrics derived from the confusion matrix.

### Human Benchmarking

Benchmarking human performance for comparison to our model was undertaken using a survey featuring a series of 25 lung ultrasound clips, sourced and labelled with agreement from 3 ultrasound fellowship trained physicians (MW, TA, RA, see Supplementary Appendix for complete survey). The survey was distributed to 100 LUS-trained acute care physicians from across Canada. Respondents were asked to identify the findings in a series of LUS loops according to the presence B lines vs normal lung (A line pattern), the characteristics of the pleural line (smooth or irregular) as well as the cause of the lung ultrasound findings (hydrostatic pulmonary edema, non-COVID pneumonia or COVID pneumonia). Responses were compared to the true, expert-defined labels consistent with our data curation process described above. Since the data used for modelling did not include normal lungs, it was decided that those four clips were discarded from analysis. Any normal diagnoses (37 of 1281 diagnoses) for the remaining clips were replaced with uniformly randomly generated diagnoses for the remaining causes.

### Explainability

We utilized the Grad-CAM method to visually explain the model’s predictions.^21^ Grad-CAM involves visualizing the gradients of the prediction of a particular image with respect to the activations of the final convolutional layer of the CNN. A heatmap is produced that is upsampled to the original image dimensions and overlaid onto the original image. The resultant heatmap highlights the areas of the input image that were most contributory to the model’s classification decision.

### Data Statement

The GitHub link to the code used to generate the DL model and the full survey data results can be found in our supplementary appendix.

### Patient and Public Involvement

Patients or the public were not involved in the design, conduct, reporting or dissemination plans of this work.

## Results

### Ultrasound Data

The data extraction process resulted in 84 cases of COVID which, as part of our effort to balance the groups for unbiased training, led to 78 of NCOVID and 81 of HPE. Further characteristics of the data are summarized in Table 2.

**Table 2:** Data profile for the 3 groups of lung ultrasound images used to train and test our model.

|  | COVID | NCOVID | HPE |
| --- | --- | --- | --- |
| Number of patients | 84 | 78 | 81 |
| Number of loops | 185 | 224 | 191 |
| Number of still images | 30419 | 44193 | 46769 |
| Female Sex (%) | 50% | 40% | 55% |
| Age (yr) | 60.6 +/- 11.3 | 56.0 +/- 16.0 | 67.2 +/- 15.3 |
| Date Range | Mar 2020 - Jun 2020 | Aug 2017 - Mar 2020 | Oct 2018 - Apr 2020 |

### Human Benchmarking

The benchmarking survey was completed by 61 physicians with a median of 3-5 years of ultrasound experience the majority of whom had done at least a full, dedicated month of ultrasound training (80.3%) and who described their comfort with LUS use as “very comfortable” (72.1%). See Supplementary Appendix for a full summary of survey data.

The results of this survey highlight that the physicians were adept at distinguishing the HPE class of B lines from COVID and NCOVID causes of B lines. For the COVID and NCOVID cases, however, significant variation and uncertainty was demonstrated. See Table 3 and “Comparing Human and Neural Networks” section below.

**Table 3:** Confusion matrices for A) the physicians (survey responses from 61 physicians classifying LUS images into their respective causes, bracketed numbers reflect classifications from the aggregated approach used to calculate AUC) and model performance on the test-2 holdback set at the B) frame and the C) encounter level.

| Physicians |  | Predicted |  |  |  |
| --- | --- | --- | --- | --- | --- |
|  |  | COVID | NCOVID | HPE | Total |
| Actual | COVID | 173 (3) | 162 (3) | 34 (2) | 369 (8) |
|  | NCOVID | 177 (4) | 163 (1) | 30 (2) | 370 (7) |
|  | HPE | 138 (0) | 102 (0) | 302 (6) | 542 (6) |
|  | Total | 488 (7) | 427 (4) | 366 (10) |  |

B
| CNN-Frames |  | Predicted |  |  |  |
| --- | --- | --- | --- | --- | --- |
|  |  | COVID | NCOVID | HPE | Total |
| Actual | COVID | 3188 | 256 | 7 | 3451 |
|  | NCOVID | 1176 | 3741 | 3 | 4920 |
|  | HPE | 109 | 1119 | 2771 | 3999 |
|  | Total | 4473 | 5116 | 2781 |  |

C
| CNN-Encounters |  | Predicted |  |  |  |
| --- | --- | --- | --- | --- | --- |
|  |  | COVID | NCOVID | HPE | Total |
| Actual | COVID | 6 | 0 | 0 | 6 |
|  | NCOVID | 1 | 6 | 0 | 7 |
|  | HPE | 0 | 3 | 4 | 7 |
|  | Total | 7 | 9 | 4 |  |

### Model Performance on Holdback Data

The model’s predictions were evaluated at both the image and encounter level. The prediction for an image is the probability vector *p* = [*p*_*COVID*_, *p*_*NCOVID*_, *p*_*HPE*_] obtained from the output of the softmax final layer, and the predicted class was taken to be *argmax*(*p*). Prediction for an encounter was considered to be 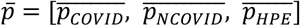, where 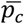 is the average predicted probability for class c over the predictions for all images within that encounter. Encounter-level predictions were computed and presented to (1) replicate the method through which real time interpretation (by clinician or machine) occurs with ultrasound by aggregating images within one or more clips to form an interpretation and (2) closely simulate a physician’s classification procedure, since the physicians who participated in our benchmarking survey were given entire clips to classify. Three models fit with our chosen architecture and set of hyperparameters were evaluated on test-1, achieving mean AUCs on the encounter level of 0.966 (COVID), 0.815 (NCOVID), and 0.902 (HPE). The model’s ultimate ability was to be determined on the 10.1% of our images that constituted the holdback data (test-2) data. On this independent data, the model demonstrated a strong ability to distinguish between the 3 relevant causes of B lines with AUCs at the encounter level of 1.0 (COVID), 0.934 (NCOVID), and 1.0 (HPE), producing an overall AUC of 0.978 for the classifier. Confusion matrices on the test-2 set at the frame and encounter level (Table 3) show strong diagonals that form the basis of these results and the performance metrics seen in Table 4.

**Table 4:**
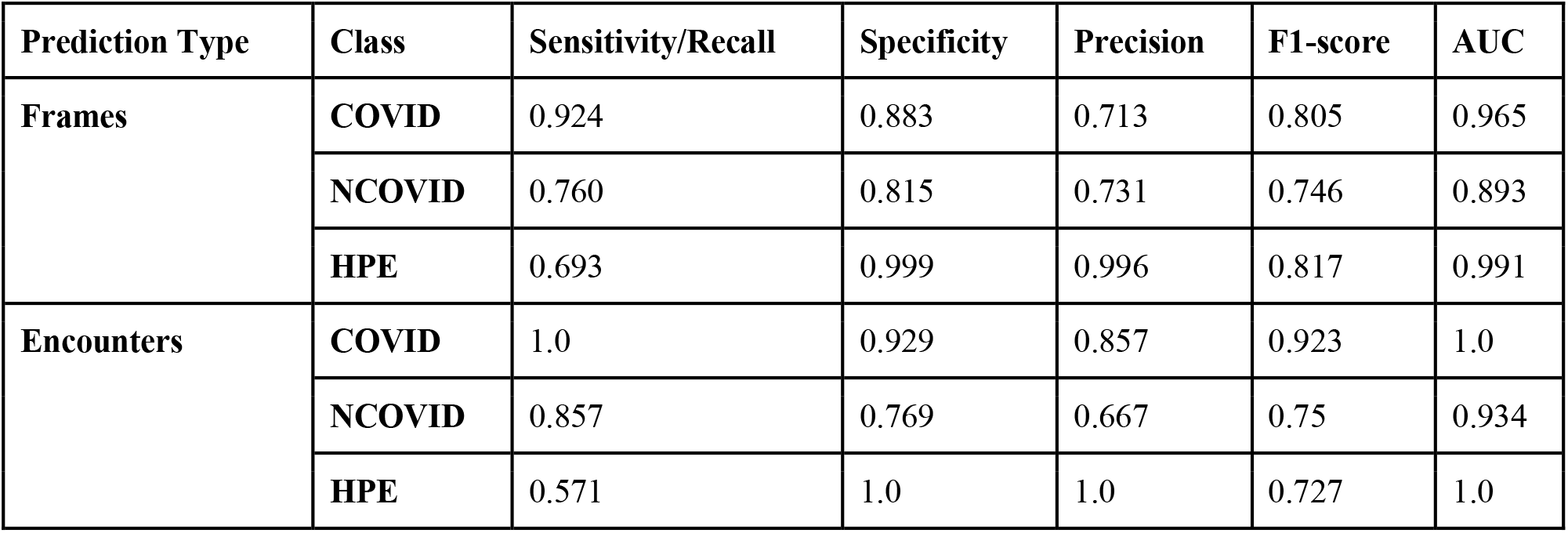
Classification performance metrics calculated from the model’s predictions and ground truth from the test-2 set. Metrics are reported at both the frame and encounter level.

| Prediction Type | Class | Sensitivity/Recall | Specificity | Precision | F1-score | AUC |
| --- | --- | --- | --- | --- | --- | --- |
| Frames | COVID | 0.924 | 0.883 | 0.713 | 0.805 | 0.965 |
|  | NCOVID | 0.760 | 0.815 | 0.731 | 0.746 | 0.893 |
|  | HPE | 0.693 | 0.999 | 0.996 | 0.817 | 0.991 |
| Encounters | COVID | 1.0 | 0.929 | 0.857 | 0.923 | 1.0 |
|  | NCOVID | 0.857 | 0.769 | 0.667 | 0.75 | 0.934 |
|  | HPE | 0.571 | 1.0 | 1.0 | 0.727 | 1.0 |

### Comparing Human and Neural Network Results

Since AUC measures a classifier’s ability to rank observations, the raw survey data (in the form of classifications, not probabilities) was processed to permit an AUC computation by considering physician-predicted probability of a LUS belonging to a specific class as the proportion of physicians that assigned the LUS to that class. The AUCs for the physicians, at face value, were 0.697 (COVID), 0.704 (NCOVID), and 0.967 (HPE), leading to an overall AUC of 0.789 (as compared to 0.978 for our model). A comparison of the human and model AUCs is graphically displayed in Figure 3. We took note of the the AUC of approximately 0.7 for the physicians when the positive class is COVID or NCOVID, as distinguishing between these classes is not known to be possible by humans. In examining the raw confusion matrix data (Table 3), this suggests near random classification (which corresponds to an AUC of 0.5) between these 2 classes - see Supplementary Appendix for a complete explanation. Given the important implications of the performance gap observed, we employed an additional step of statistical validation for our findings through a Monte Carlo Simulation (MCS, see Supplementary Appendix for full details) of human performance, based on our survey results, across one million exposures to our test-2 data.^22^ After simulating this performance one million times, the MCS yielded an average AUC of 0.840 across all three classes, with very few cases matching or exceeding the performance of the CNN. Thus, we can conclude that our model exceeds human performance, and in particular that the model can distinguish between COVID and NCOVID (p < 0.01).

**Figure 3:**
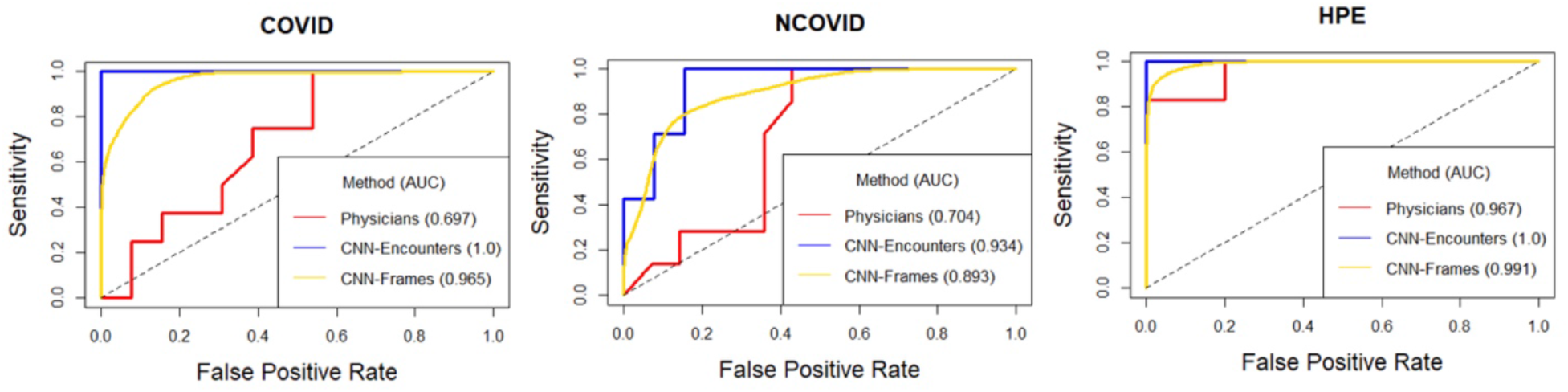
Receiver operating characteristic curves across the three classes of images that our human benchmarking (physicians) and our model (convolutional neural network: CNN) were tasked with interpreting. The model’s performance on the test-2 (holdback) image set is plotted for both individual images and across the entire image set from one encounter. In all image categories, it can be seen that the model interpretation accuracy exceeded that of the human interpretation.

### Explainability Results

The Grad-CAM explainability algorithm was applied to the output from the model on the holdback data. The results are conveyed by color on the heatmap, overlaid on the test-2 input images. Blue and red regions correspond to highest and lowest prediction importance respectively. As the results seen in Figure 4 demonstrate, the key activation areas for all classes were centered around the pleura and the pleural line.

**Figure 4:**
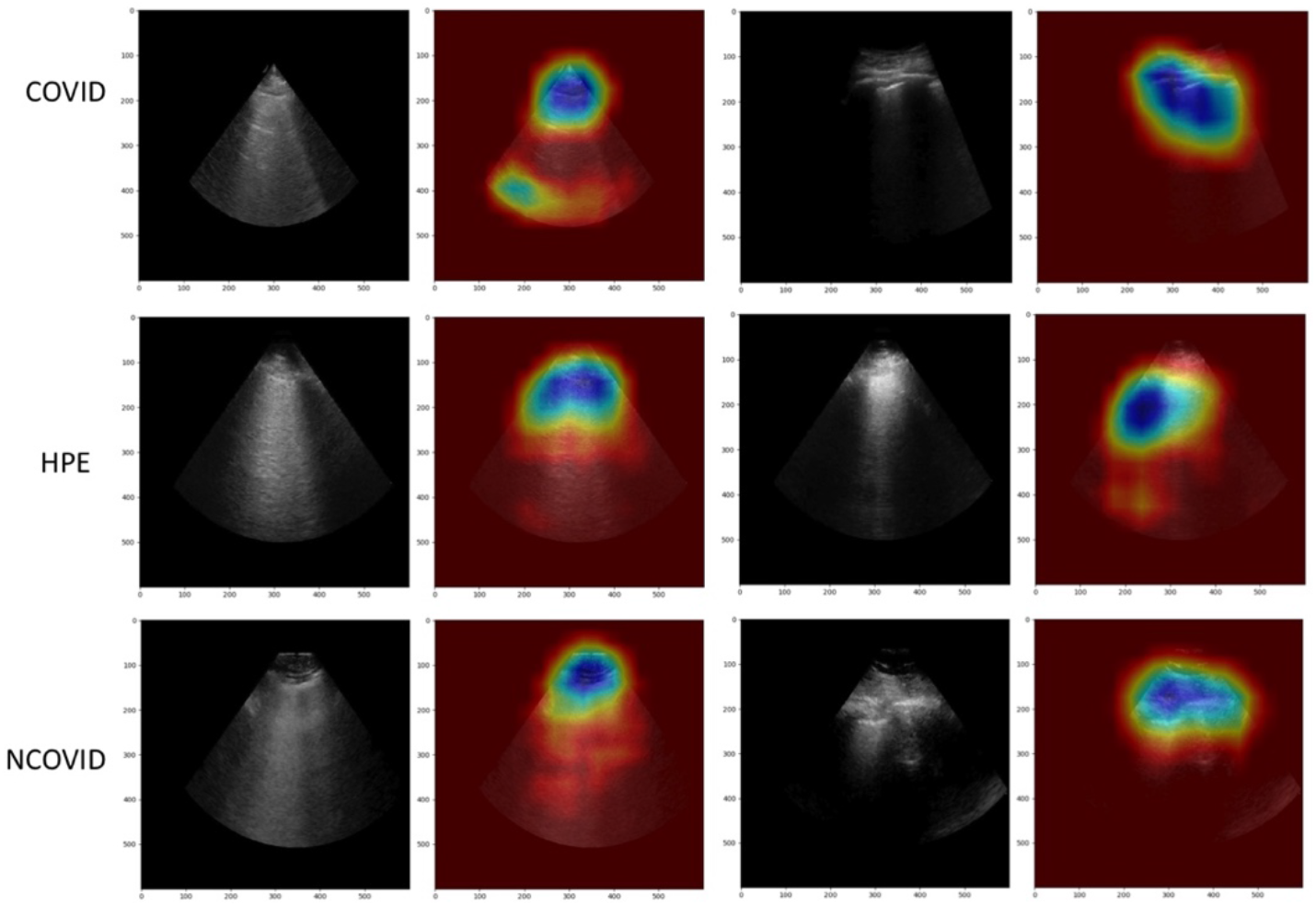
Grad-CAM heatmaps corresponding to a selection of our model’s predictions. Blue areas reflect the regions of the image with the highest contribution to the resulting class predicted by the model. In all cases, the immediate area surrounding the pleura appears most activated. COVID: COVID-19 pneumonia, HPE: hydrostatic pulmonary edema, NCOVID: non-COVID related acute respiratory distress syndrome.

## Discussion

In this study, a deep learning model was successfully trained to distinguish the underlying pathology in similar point-of-care lung ultrasound images containing B lines. The model was able to distinguish COVID-19 from other causes of B lines and outperformed ultrasound trained clinician benchmarks across all categories. Our results are the first of their kind to support that digital biomarker profiles may exist within lung ultrasound images.

Our model was developed using a data set of 243 patients (600 video loops/121,381 frames) which is modest by machine learning standards. Owing to the scarcity of labelled lung ultrasound data, this data volume does compare favorably to other published lung ultrasound work.^23–25^ Given the implications of successfully classifying LUS images, it was essential for us to protect against overfitting. While many approaches exist to avoid an overfit model, we, in addition to multiple data augmentation techniques, reserved 10% of our data (test-2) as a hold-back set, not involved in model fitting or selection. This approach mimics the unbiased, generalizable performance desired of an image classifier and is familiar to other notable deep learning vision research in medicine.^9–11^

Deep learning has shown similarly favorable results in recent CXR and CT studies of COVID-19.^26,27^ Given LUS image creation is fundamentally different (producing artifacts, rather than anatomic images of the lung), it could not be expected that our work with LUS would have yielded such similar results. The value of identifying such accuracy in a LUS model rests in the ability of LUS (unlike CT or CXR) to be delivered by limited personnel, at low cost and in any location.

Lung ultrasound artifact analysis has existed for several years in some commercially available ultrasound systems and has also been described using various methods in the literature.^23,28,29^ Automating the detection of canonical findings of LUS, these techniques are convenient and serve to achieve what clinicians may be trained to do with minimal training.^30^ With attention to COVID-19, LUS has been shown to inform clinical course and outcome^31^, creating some further momentum toward broader LUS competence. As our work opens the door toward plausible early, automated COVID identification using LUS, DL techniques to auto-generate clinical severity score for COVID has also recently been described.^24^ The eventual integration of various DL models into ultrasound hardware seems plausible as a method to achieve real time, point-of-care diagnosis and prognosis of COVID or other specific respiratory illnesses.

The implications of our work, at time of writing, are strongly attached to the current challenges and importance of COVID-19 diagnosis. Our results point to a unique, pixel-level signature within the COVID-19 image. Though the exact mechanism of distinction is unknown, the heat map results suggest that subvisible variations in the pleural line itself is most active in driving the model’s performance. The precise taxonomical implications of our findings, whether they are driven by COVID-19, coronaviruses or viruses a whole, will require additional research.

Our study has some important limitations. The first relates to the opaqueness that is implicit to deep neural networks. Despite using Grad-CAM, the decisions by the trained model are not outwardly justified and we are unable to critique its methods and must trust its predictions. Our benchmarking survey did not exactly replicate the questions posed to our neural network which made our statistical analysis more complex than it might have needed to be otherwise. The other limitations of our study related to the data size and sources. Though our model performance signal was strong, the addition of further training data can only aid with generalizability of the model. Future work will focus on validation through multi-center collation of COVID LUS encounters. Lastly, our data were all from hospitalized patients and our results may not generalize to those who are less ill.

## Conclusions

With strong performance in distinguishing lung ultrasound images of COVID-19 from mimicking pathologies, a trained neural network exceeded human interpretation ability and raises the possibility of disease-specific, subvisible features contained within lung ultrasound images. Further research using well-labelled, multi-center data is indicated.

## Data Availability

https://github.com/bvanberl/covid-us-ml

## Author Contributions

All authors were involved in the authorship of the manuscript, figures and tables. Overall project design and oversight (RA, BV), data management (TA, DW, RA, JH, MW), survey creation and distribution (MW), model training (BV, DW, NP), statistical analysis (NP), figure generation (JH, NP, BV) and literature search (RC, RA).

## Acknowledgments

The authors would like to acknowledge the computational and technical support from CENGN (Canada’s Centre of Excellence in Next Generation Networks), Mr. Matt Ross from the City of London, Mrs. Kristine Van Arsen from the Division of Emergency Medicine and the clinician-sonographers at London Health Sciences Centre who faithfully record and annotate their lung ultrasound studies.

## Supplementary Methods

### Image Beam Isolator Method

In order to remove extraneous information, an algorithm was written to isolate the ultrasound beam from the rest of the image by mask. This algorithm was designed to be generalizable to ultrasound beams of all widths and positions as different machines have different interfaces. To allow the isolation algorithm to find the best quality mask, the largest continuous contour was detected using classical computer vision techniques for every frame in a video, and the frame with the largest contour was used for calculations of the ultrasound beam. The two linear edges were derived by finding every point on the contour that was both the top-most and left- or right-most in every column and row, and finding two lines of best fit for both sets of points. The bottom circular edge was then calculated using the intersection of these two lines as the vertex, and fitted to all the bottom-most points on the contour. A mask was generated using these edges and subsequently applied to every frame in the video, as all frames in the same video had the beam in the same position. If the algorithm was unable to find a matching ultrasound beam within empirically determined limits due to poor-quality beams, it reverted to using the contour as the mask. This technique was successful at removing information outside of the ultrasound beam and preserving information that may have been left out by the contour; unfortunately, text or interface artifacts contained within the beam (a very uncommon occurrence) were retained by the algorithm. Figure S1 depicts this process.

### Model Architecture Selection

Figure S2 provides an overview of our process for this phase and the model validation phase. Transfer learning (TL) is a technique that utilizes previously learned weights from a neural network trained on a separate problem and has previously shown success with CNNs for other ultrasound classification problems. Initial TL trials involved fine-tuning common architectures that included ResNet50V2, ResNet101V2, VGG16, InceptionV3, InceptionResNetV2, MobileNetV2 and Xception. After iterating through these architectures using training experiments, Xception achieved highest performance on different validation subsets and on the test-1 set. The model’s weights were initialized with weights originating from an Xception model that was pre-trained on ImageNet<u>^19^</u>, which are freely available via TensorFlow. Pre-trained weights from the fully connected layer of TensorFlow’s Xception model were left out, as we investigated different options for fully connected layers that follow the convolutional layers. Multiple sets of hyperparameters were trialled.

### Model Performance: Prediction Method

The prediction for an image is the probability vector *p* = [*p*_*COVID*_, *p*_*NCOVID*_, *p*_*HPE*_] obtained from the output of the softmax final layer, and the predicted class was taken to be *argmax*(*p*). Prediction for an encounter was considered to be 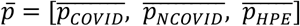, where 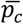 is the average predicted probability for class c over the predictions for all images within that encounter.

### Model Training, Code and Hardware

The CNN was trained using the Adam optimizer with a learning rate (*α*) of 1 × 10^−6^ to minimize the binary cross-entropy loss function. The magnitude of the loss function for any particular prediction during training was weighted by the representation in the dataset of each class. We employed early stopping with a patience of 3 epochs, conditioned on validation loss, as a means of regularization.

All code was written in Python 3.7. The TensorFlow open source machine learning platform (www.tensorflow.org) was used to define and train the CNN model. Our GitHub repository contains the code used to generate the results presented in this paper and can be seen at https://github.com/bvanberl/covid-us-ml.

In-house hardware used for training experiments included a personal computer running Windows 10, equipped with an Intel^®^ Core i9-9900K^®^ processor at 3.6 GHz and a NVIDIA^®^ GeForce^®^ RTX 2080 Ti GPU with 11GB of memory. Three virtual machines (VMs) were also employed for training experiments. Each VM was running Ubuntu 18.04 LTS Server. The VMs had access to NVIDIA^®^ Tesla T4 GPUs, each with 16GB of memory and novel Tensor Cores. One VM had 5 such GPUs and the others had 3. Care was taken to optimize the use of the Tensor Cores on the T4 GPUs by setting the data types of some variables in the neural network model to be 16-bit floating-point, thereby further accelerating training experiments.

### Model Validation Methods

Expanding on the validation methods in our manuscript. At the start of each training experiment, the training set was randomly split into a training subset and a validation subset. As a result, the validation subset differed in each successive experiment. Since early stopping was employed, the validation subset was inherently involved in the model selection process. After each training experiment, the model was assessed based on its performance on the validation subset and the test-1 set. In this way, test-1 was used for model selection with the dynamic validation subsets. Once settled on a model architecture with a particular set of hyperparameters, we entered the model validation phase. To train the final model, we combined the training set and test-1 to form a larger training set, which was subsequently split into a training and validation subset for the final experiment. Once training was complete, the trained model was evaluated by calculating performance metrics from its predictions on the examples in the test-2 set.

### Human/Machine comparison and Monte Carlo Simulation

Since AUC measures a classifier’s ability to rank observations, the raw survey data (in the form of classifications, not probabilities) was processed to permit an AUC computation by considering physician-predicted probability of a LUS belonging to a specific class as the proportion of physicians that assigned the LUS to that class.

Although all three AUCs obtained by the physicians reached the 0.7 threshold associated with acceptable discrimination, the ROC curves and their associated scores actually suggest that physicians have very limited to no ability in distinguishing between COVID and NCOVID. All three AUCs were driven by the physicians’ ability to separate HPE from the other two classes. When the positive class is either COVID or NCOVID, the top right part of the ROC curve clearly exceeds random performance. The high level of performance associated with this section occured because very few physicians diagnosed COVID/NCOVID ultrasounds as HPE. Consequently, even with a low threshold probability, HPE ultrasounds were accurately identified as part of the negative class, reducing the false positive rate while the sensitivity remained high. This section of the curve inflated the AUC above random performance. The physicians’ ability to distinguish between COVID and NCOVID is better represented by the bottom left part of the ROC curve. When either COVID or NCOVID is the positive class, the bottom part of the curve approximately follows along the 45° line associated with random performance.

We used two different approaches to adjust the survey data in order to quantitatively validate the above qualitative observations. Both were designed to remove HPE, converting the problem to a binary (COVID vs NCOVID) problem. The first approach was the same as the one described in the Human Benchmarking section, where physician diagnoses that were no longer relevant (in this case, HPE) were removed and replaced with randomly generated diagnoses from the remaining candidates. The second approach conditioned the physicians’ aggregate diagnosis on the knowledge that the ultrasound was not HPE. The AUCs obtained from these two procedures were 0.455 and 0.429 respectively, both below the 0.5 threshold associated with random performance.

In order to perform a statistical test comparing the model’s results to human performance, and in particular to test if the model was able to distinguish between COVID and NCOVID, we performed a Monte Carlo Simulation (MCS). For the MCS, we used a classifier that perfectly separated HPE from COVID and NCOVID (i.e., all HPE cases were predicted as HPE with a probability of one and all COVID/NCOVID cases were assigned zero probability of being HPE), but guessed when deciding between COVID and NCOVID. For the COVID and NCOVID cases, the predicted probability of the observation belonging to the COVID class was drawn from a Uniform(0, 1) distribution and the remaining probability was assigned to the NCOVID class. This classifier provided an upper bound on performance for a model unable to distinguish between COVID and NCOVID (i.e., an upper bound on human performance). Thus, if a model is shown to outperform this model, it must be able to distinguish between COVID and NCOVID. We simulated the performance of this classifier on our testing dataset one million times in order to obtain the distribution of its overall AUC. The results are shown in **Figure S3**, with the dashed line representing the AUC of the CNN. The unusual distribution was caused by a correlation of 1.0 between the AUCs when COVID and NCOVID were the positive class. The results indicate that the model outperforms this classifier (p < 0.01). Therefore, we can comfortably assert that the model outperforms humans and can distinguish between COVID and NCOVID.

## Supplemental Tables

**Table S1:** Demographic information of participants in human benchmarking survey

|  | Total number (%) |
| --- | --- |
| **Total number of participants** | 61 |
| ***Level of Training*** |  |
| Attending Physician | 28 (45.9) |
| Sr. Resident | 33 (54.1) |
| ***Experience with POCUS (yrs)*** |  |
| <1 | 3 (4.9) |
| 1-3 | 23 (37.7) |
| 3-5 | 22 (36.1) |
| 5-10 | 7 (11.5) |
| >10 | 6 (9.8) |
| ***Specific POCUS training*** |  |
| None | 0 |
| POCUS fellowship | 14 (23) |
| POCUS residency elective | 49 (80.3) |
| Live POCUS course | 43 (70.5) |
| Online POCUS course | 11 (18) |
| ***Comfort with lung ultrasound (LUS)*** |  |
| Very comfortable, routine use | 44 (72.1) |
| Somewhat comfortable | 16 (26.2) |
| Limited comfort | 1 (1.6%) |
| Not comfortable with use | 0 |
| ***Specific applications of LUS used*** |  |
| Pneumothorax | 54 (88.5) |
| Pleural effusion | 61 (100) |
| Interstitial syndrome | 61 (100) |
| Consolidation (Pneumonia/atelectasis) | 52 (85.2) |
| ***Comfort in distinguishing hydrostatic pulmonary edema versus infectious/inflammatory etiologies*** |  |
| Yes | 45 (73.8) |
| No | 16 (26.2) |

## Supplemental Figures

**Figure S1:**
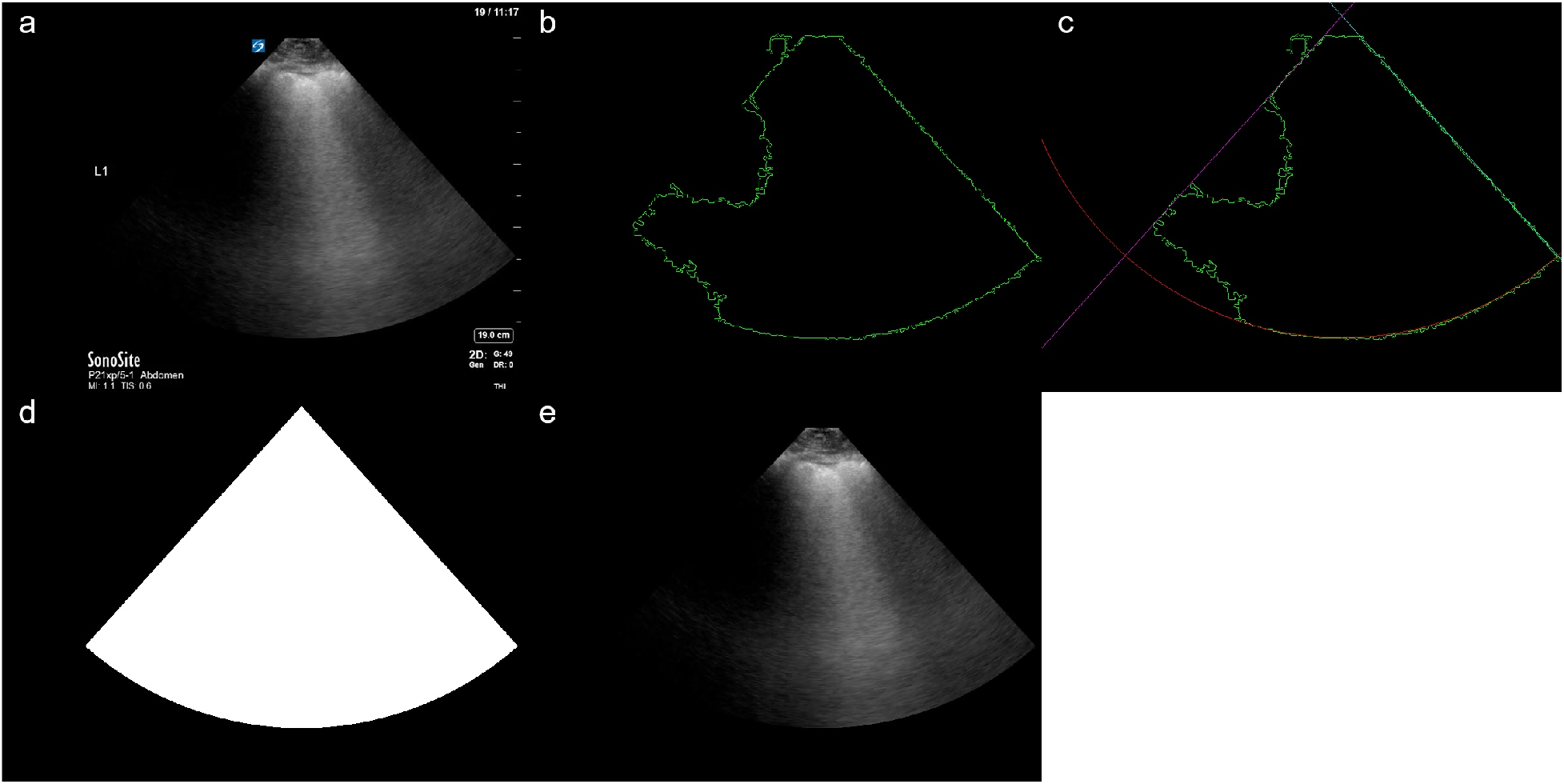
Processing of one frame of an ultrasound clip. a) the original frame, b) the contour of the ultrasound beam, c) the edges as calculated by the algorithm, d) the mask generated by the edges, and e) the final processed frame.

**Figure S2:**
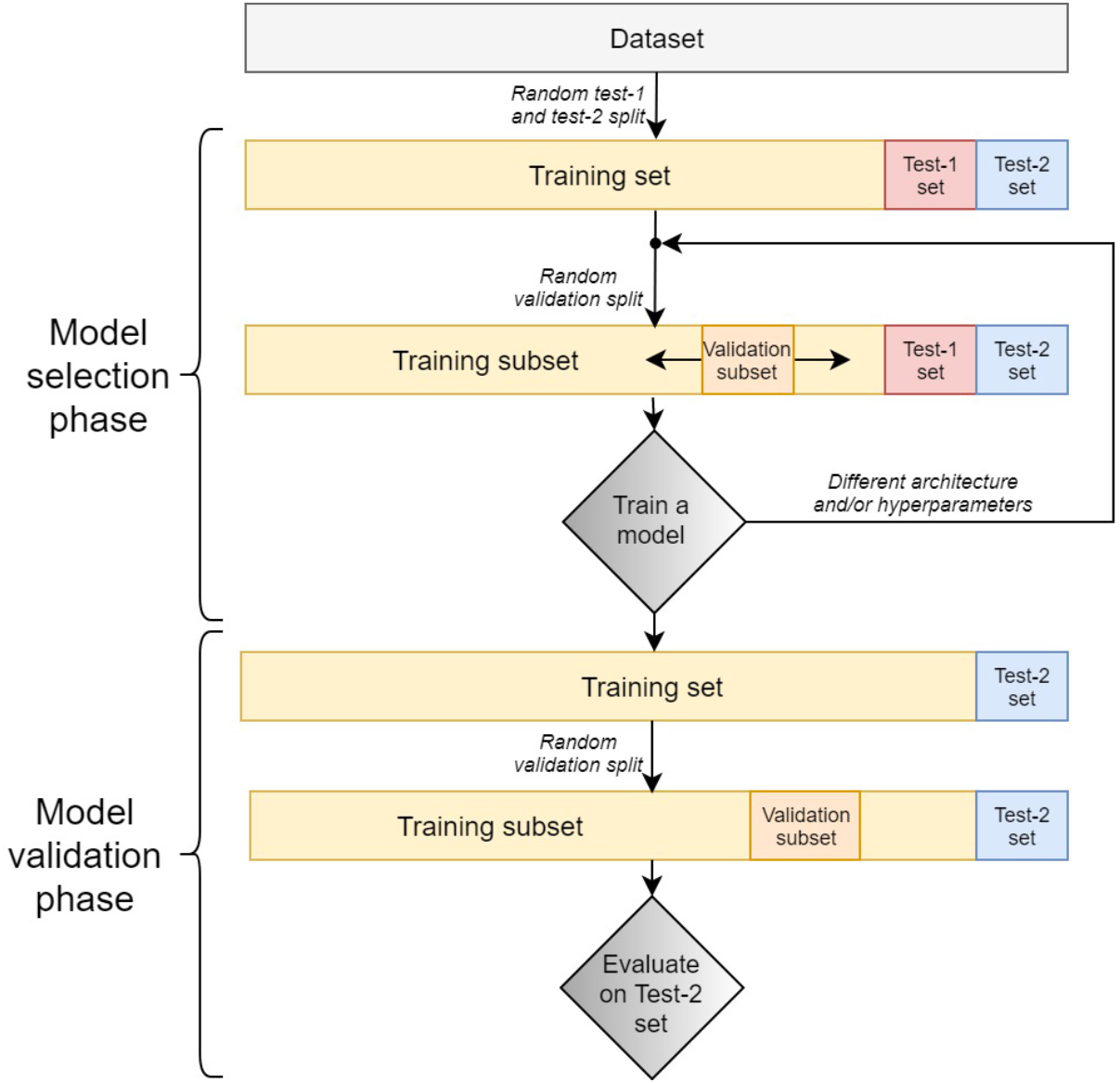
Workflow applied for the model selection and model validation phase of this project.

**Figure S3:**
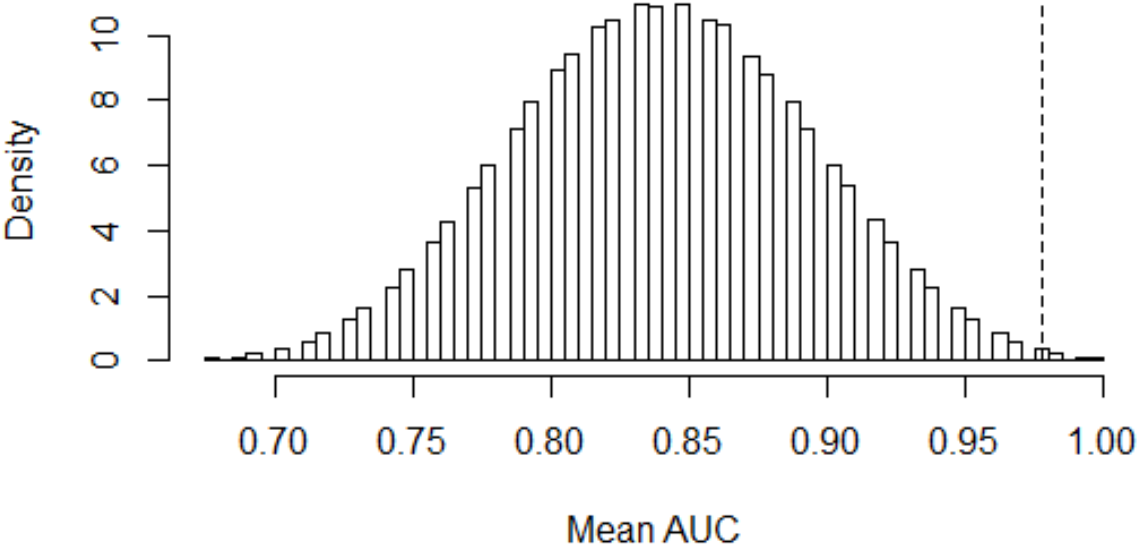
A histogram displaying the results of the MCS with one million runs. The vertical dashed line represents the performance of the neural network.

